# CBD use characteristics associated with perceived effects and satisfaction among CBD product users in Japan: an exploratory analysis of a cross-sectional web-based survey

**DOI:** 10.64898/2026.10.12.26365299

**Authors:** Aya Kuroda, Chenyu Pan, Lyie Nitta, Chihiro Nozaki

**Author notes:** Correspondence: Chihiro Nozaki, Wakamatsu-cho 2-2, Shinjuku-ku, Tokyo 162-8480, Japan.

## Abstract

**Background:** Cannabidiol (CBD) products are sold in many forms for many purposes, but it is largely unknown which characteristics of real-world use relate to how users evaluate their experience.

**Methods:** We analyzed an anonymous web-based survey of 377 CBD product users in Japan. We evaluated perceived-effect (0–5) and satisfaction (0–6) scores using main regression models of use frequency adjusted for age, sex, and body mass index. Separate supplementary models examined estimated daily CBD amount and other-cannabinoid co-use, purposes of use, or specific CBD amount-reporting status.

**Results:** Use frequency was positively associated with perceived effects (B = 0.148, 95% CI 0.091 to 0.205) and satisfaction (B = 0.113, 95% CI 0.049 to 0.177; both p < 0.001).

Estimated daily amount showed no clear association with either outcome after adjustment, whereas participants who reported specific milligram information had higher scores on both (B = 0.543, p < 0.001 and B = 0.559, p = 0.002). Amount-reporting status was associated with product form (tincture 48.5% vs 30.8%; smoking or inhalation 58.5% vs 75.7%; both FDR-adjusted p = 0.009), whereas no statistically significant group differences were found for age, sex, body mass index, use frequency, or sleep timing. Purposes of use and sleep-timing indicators showed no clear associations with the outcomes.

**Conclusions:** Subjective evaluation of CBD use was more consistently related to how often CBD was used and whether a specific milligram amount was reported than to how much was reportedly taken, and amount-reporting status was related to product form. Specific CBD amount-reporting status may be a useful complementary dimension for characterizing real-world CBD use.

## Introduction

Cannabidiol (CBD) products are available in Japan in a wide range of forms, including oils, tablets, smoking or inhalation products, edibles, and topical preparations (Ministry of Health, Labour and Welfare, 2021; Masataka et al., 2022). Surveys conducted outside Japan have shown that CBD is used for diverse purposes, including sleep, relief of anxiety symptoms, stress reduction, and relaxation (Moltke and Hindocha, 2021; Wheeler et al., 2020).

The legal framework for cannabinoid products in Japan has also changed in recent years (Ministry of Health, Labour and Welfare, 2024). Against this background, characterizing how commercially available CBD products are used in Japan, by whom, for what purposes, and in what forms, is increasingly important. Japanese data on CBD use nevertheless remain limited (Masataka et al., 2022), and few studies have examined which use characteristics are associated with perceived effects and satisfaction when multiple characteristics and demographic factors are considered together.

Fortin et al. (2025) used hierarchical classification to identify distinct profiles among people who co-used cannabis and CBD, treating users as a heterogeneous population characterized by combinations of multiple features. Comparable evidence is scarce for CBD product users in Japan, particularly when use frequency, amount-reporting, co-use of other cannabinoids, purposes of use, and product forms are considered jointly.

One of the most commonly reported purposes of CBD use is sleep improvement (Moltke and Hindocha, 2021; Masataka et al., 2022), but this purpose is conceptually distinct from a user’s own sleep timing. Individual sleep timing can be characterized by established indices such as sleep-corrected mid-sleep on free days (MSFsc) and social jetlag (SJL) (Roenneberg et al., 2003; Wittmann et al., 2006).

We therefore analyzed data from an anonymous web-based survey of CBD product users in Japan conducted by our group. We described respondents’ basic characteristics, purposes of use, product forms, use frequency, estimated daily CBD amount, specific CBD amount-reporting status, and co-use of other cannabinoids, and examined how these characteristics, together with MSFsc and SJL, were associated with perceived effects and satisfaction.

## Methods

### Study Design, Participants, and Ethics

This cross-sectional, anonymous web-based survey of CBD product users in Japan was conducted between 24 November 2022 and 5 January 2023. Respondents were recruited at a CBD-related event, through posters at CBD retail stores, and via social media (Twitter, Facebook, Instagram). Participation was voluntary, and consent to research use of responses was obtained at the time of response. Waseda University’s ethics review committee for research with human subjects determined that ethical review was not required (reference no. 2023-HN001). Of 382 respondents, five were excluded: four whose current CBD use could not be confirmed, and one who asked that their responses not be analyzed. The analytic sample therefore comprised 377 participants.

### Measures

The outcomes were single-item scores for perceived effects (0–5) and satisfaction (0–6). Higher scores indicated stronger perceived effects or greater satisfaction, and both were analyzed as continuous variables.

CBD use frequency was converted to days of use per week, and estimated daily CBD amount to a representative value for each reported milligram range. Specific CBD amount-reporting status indicated whether a respondent reported specific milligram information in the daily-amount item. Other explanatory variables were co-use of cannabinoids other than CBD, four non-exclusive purpose-of-use categories (sleep-related, psychological or stress-related, pain-related, and relaxation-related), and seven product forms. Age (four categories), sex, and BMI were included as covariates.

MSFsc and SJL were calculated from reported bedtimes and wake times on usual days (for example, weekdays) and on days off with no scheduled activities (Roenneberg et al., 2003; Wittmann et al., 2006). Responses suggesting 12-hour-format entry errors were corrected using a uniform rule, and daytime sleep schedules were retained. Survey items, full definitions, and coding rules are given in Supplementary Methods and Supplementary Tables S1 to S3.

### Statistical Analysis

Each outcome was analyzed by ordinary least squares regression with HC3 robust standard errors, using complete cases. The main model included CBD use frequency, age category, sex, and BMI. Supplementary models added estimated daily CBD amount and co-use of other cannabinoids (model A), the four purpose-of-use categories (model B), or specific CBD amount-reporting status (model C). MSFsc and SJL were each examined in separate models adjusted for use frequency, specific CBD amount-reporting status, age category, sex, and BMI.

Participants who did and did not report a specific CBD amount were compared using chi-square tests (Cramér’s V) and Mann–Whitney U tests (rank-biserial correlation), with Benjamini–Hochberg false discovery rate (FDR) correction across the seven product forms. Unadjusted associations were examined by Spearman rank correlation with FDR correction, and patterns of use were explored by Ward’s hierarchical clustering and principal component analysis. Model diagnostics and sensitivity analyses are described in Supplementary Methods. Analyses were performed in Python 3.13.5 (pandas 2.2.3, NumPy 2.3.5, SciPy 1.17.0, statsmodels 0.14.6, and scikit-learn 1.8.0). All tests were two-sided, with a significance level of 0.05.

## Results

### Participant Characteristics

Perceived-effect scores were available for 373 participants and satisfaction scores for all 377 (Table 1). Median CBD use frequency was 3.5 days/week [1.5–7.0]. A specific milligram amount was reported by 270 participants (71.6%); among the 268 whose daily amount could be quantified, the median was 18.0 mg [5.0–38.0]. Co-use of other cannabinoids was reported by 182 (48.3%). The most frequently reported product forms were smoking or inhalation products (63.4%) and tinctures (43.5%). Notably, 107 participants (28.4%) did not provide specific milligram information.

**Table 1.**
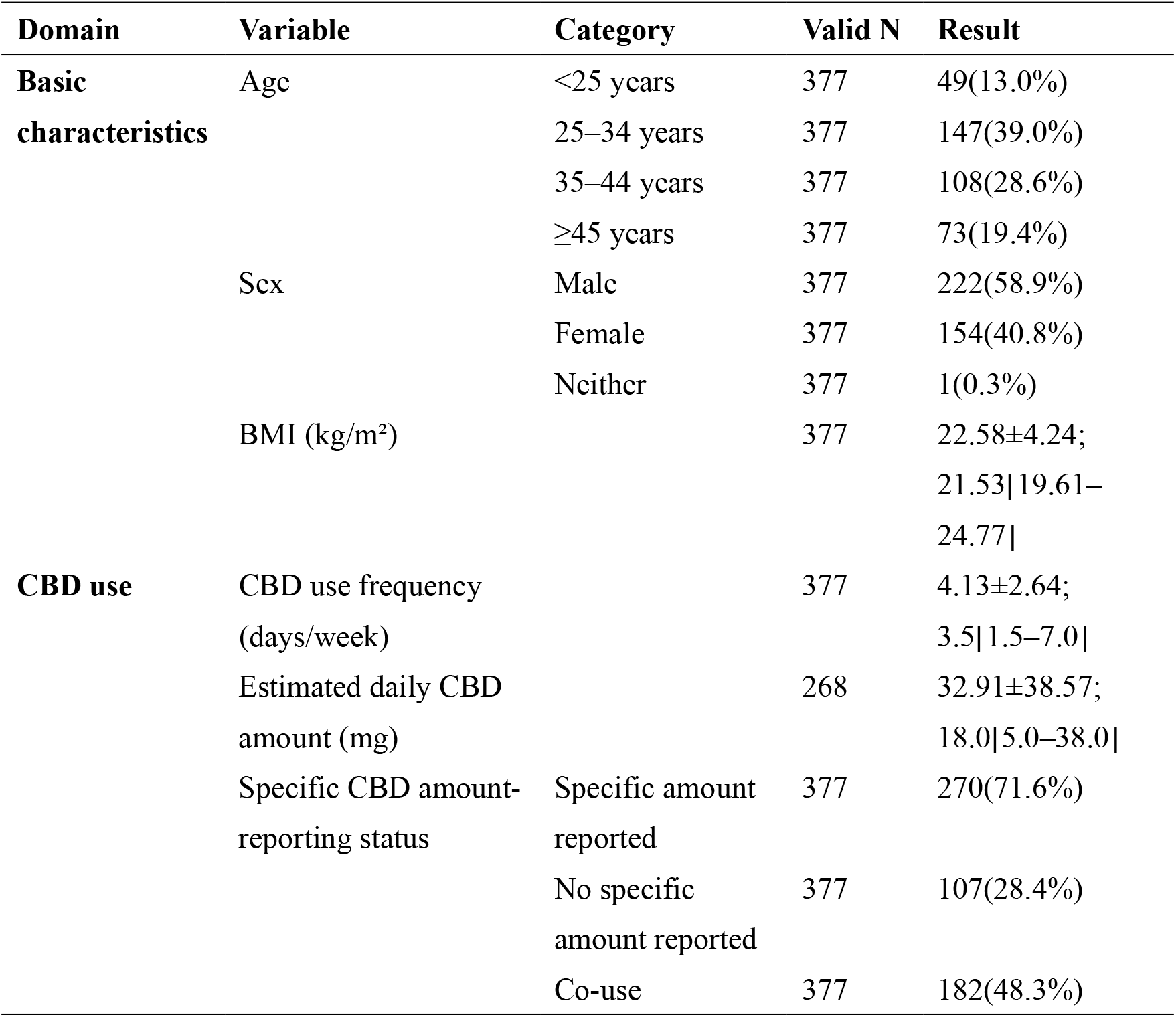
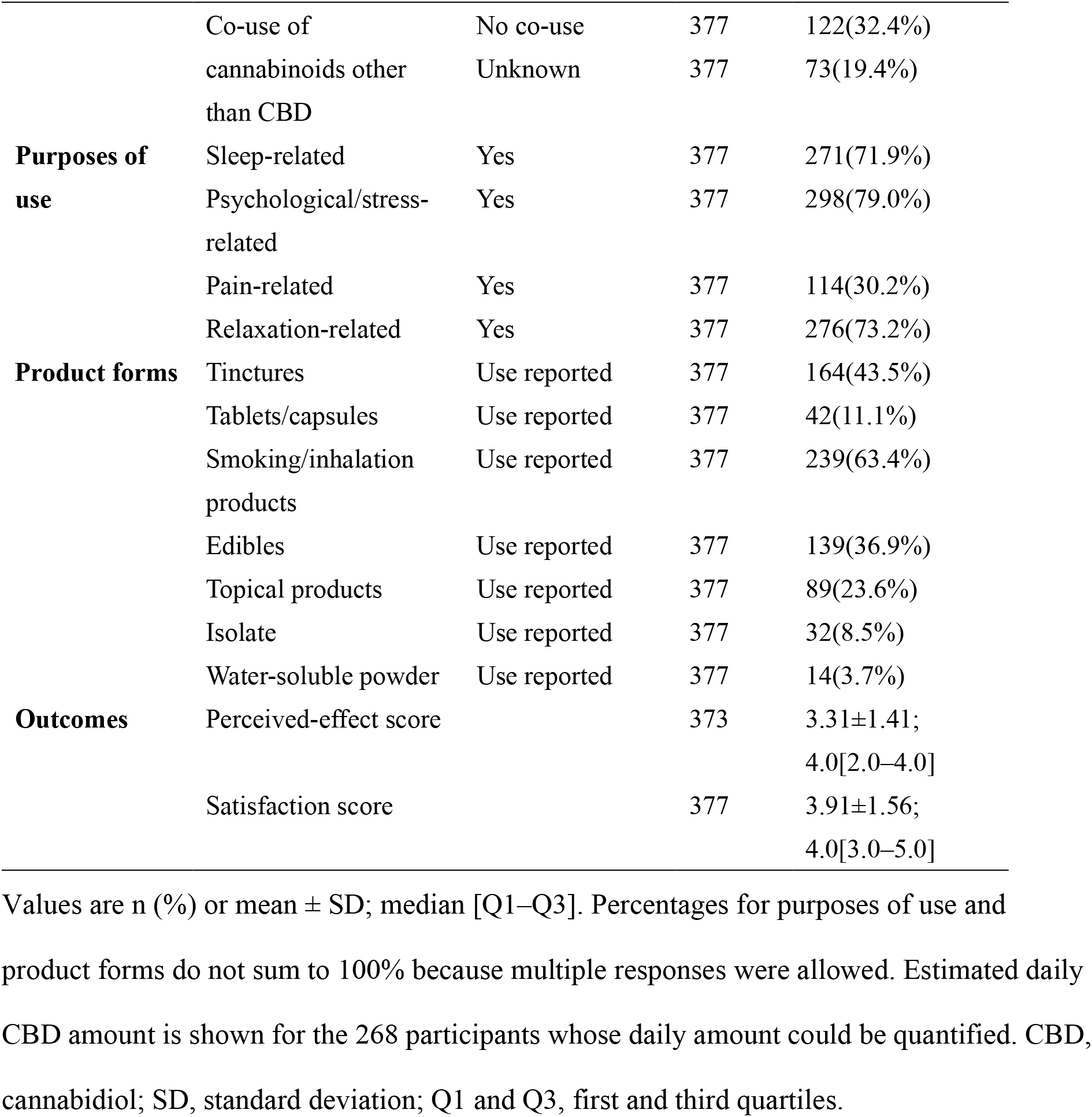
Participant characteristics, CBD use, purposes of use, product forms, and outcomes.

### Use frequency, estimated amount, and amount-reporting status

In the main model, CBD use frequency was positively associated with both perceived-effect and satisfaction scores (Table 2; adjusted R-squared 0.074 and 0.043, respectively). This association remained when frequency was analyzed as five categories (Supplementary Table S5) and in all three supplementary models. After adjustment, estimated daily CBD amount was not clearly associated with perceived-effect score (B = 0.0035 per mg, 95% CI −0.0004 to 0.0075, p = 0.081) or satisfaction score (B = −0.0013, 95% CI −0.0070 to 0.0045, p = 0.665; model A), and this did not change when the representative value for the highest amount category was varied (Supplementary Table S4). No clear adjusted associations with either outcome were detected for co-use of other cannabinoids (model A) or the four purpose-of-use categories (model B). In contrast, participants who reported a specific CBD amount had higher perceived-effect scores (B = 0.543, 95% CI 0.223 to 0.864, p < 0.001) and satisfaction scores (B = 0.559, 95% CI 0.214 to 0.903, p = 0.002) than those who did not (model C). Full coefficients for all models are given in Supplementary Table S4.

**Table 2.**
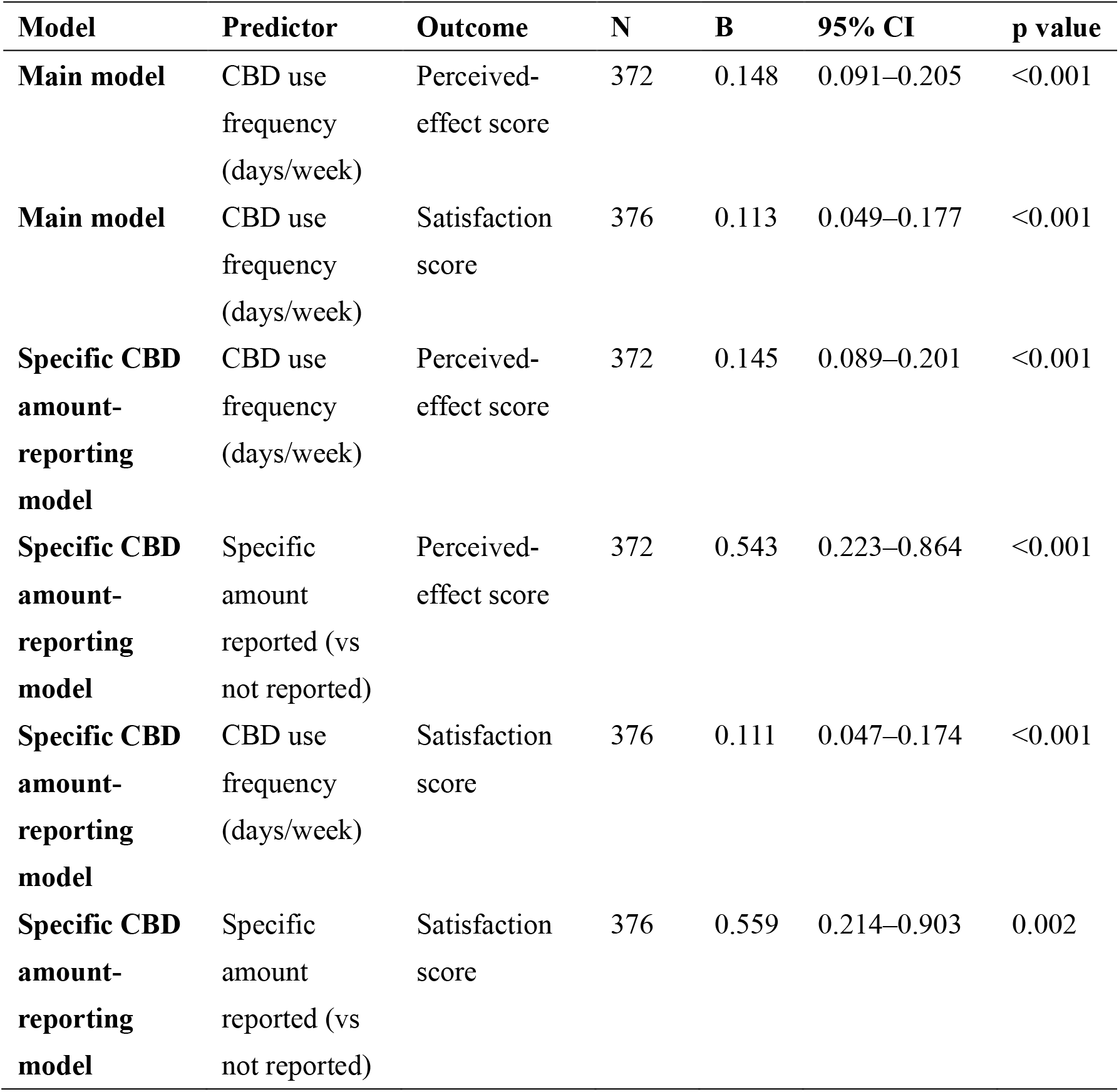
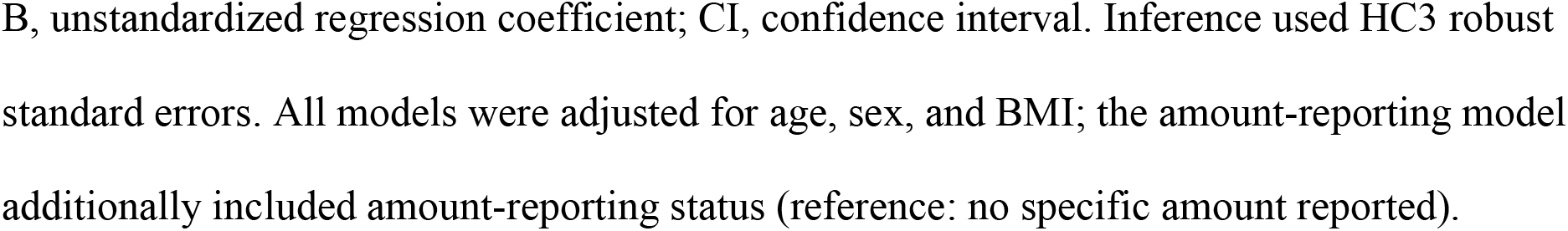
Principal adjusted associations.

| **Model** | **Predictor** | **Outcome** | **N** | **B** | **95% CI** | **p value** |
| --- | --- | --- | --- | --- | --- | --- |
| **Main model** | CBD use frequency (days/week) | Perceived-effect score | 372 | 0.148 | 0.091–0.205 | <0.001 |
| **Main model** | CBD use frequency (days/week) | Satisfaction score | 376 | 0.113 | 0.049–0.177 | <0.001 |
| **Specific CBD amount-reporting model** | CBD use frequency (days/week) | Perceived-effect score | 372 | 0.145 | 0.089–0.201 | <0.001 |
| **Specific CBD amount-reporting model** | Specific amount reported (vs not reported) | Perceived-effect score | 372 | 0.543 | 0.223–0.864 | <0.001 |
| **Specific CBD amount-reporting model** | CBD use frequency (days/week) | Satisfaction score | 376 | 0.111 | 0.047–0.174 | <0.001 |
| **Specific CBD amount-reporting model** | Specific amount reported (vs not reported) | Satisfaction score | 376 | 0.559 | 0.214–0.903 | 0.002 |

B, unstandardized regression coefficient; CI, confidence interval. Inference used HC3 robust standard errors. All models were adjusted for age, sex, and BMI; the amount-reporting model additionally included amount-reporting status (reference: no specific amount reported).

### Characteristics associated with amount-reporting status

Participants who did and did not report a specific CBD amount did not differ in age (p = 0.981), sex (p = 0.613), BMI (p = 0.780), CBD use frequency (p = 0.560), or MSFsc (p = 0.339), but both outcome scores were higher among those who reported a specific amount in unadjusted rank-based comparisons (rank-biserial correlation 0.213 and 0.227, both p < 0.001). The groups also differed in product form: tincture use was more common among those who reported a specific amount (48.5% vs 30.8%, FDR-adjusted p = 0.009) and smoking or inhalation product use less common (58.5% vs 75.7%, FDR-adjusted p = 0.009), whereas the other five product forms did not differ after FDR correction (Supplementary Table S6).

Amount-reporting status therefore did not differ by background characteristics but did differ by product form.

### Unadjusted correlations

Perceived-effect and satisfaction scores were strongly correlated (rho = 0.681). After FDR correction, CBD use frequency (rho = 0.277 and 0.193) and amount-reporting status (rho = 0.172 and 0.181) were the only use characteristics showing FDR-significant correlations with both outcomes (Figure 1). Perceived-effect score was also correlated with estimated daily CBD amount (rho = 0.163), co-use of other cannabinoids (rho = 0.193), pain-related purpose (rho = 0.150), and tablet or capsule use (rho = 0.175), and satisfaction score with tincture use (rho = 0.158; Supplementary Table S7). These were thus the same two characteristics that were associated with both outcomes after adjustment.

**Figure 1.**
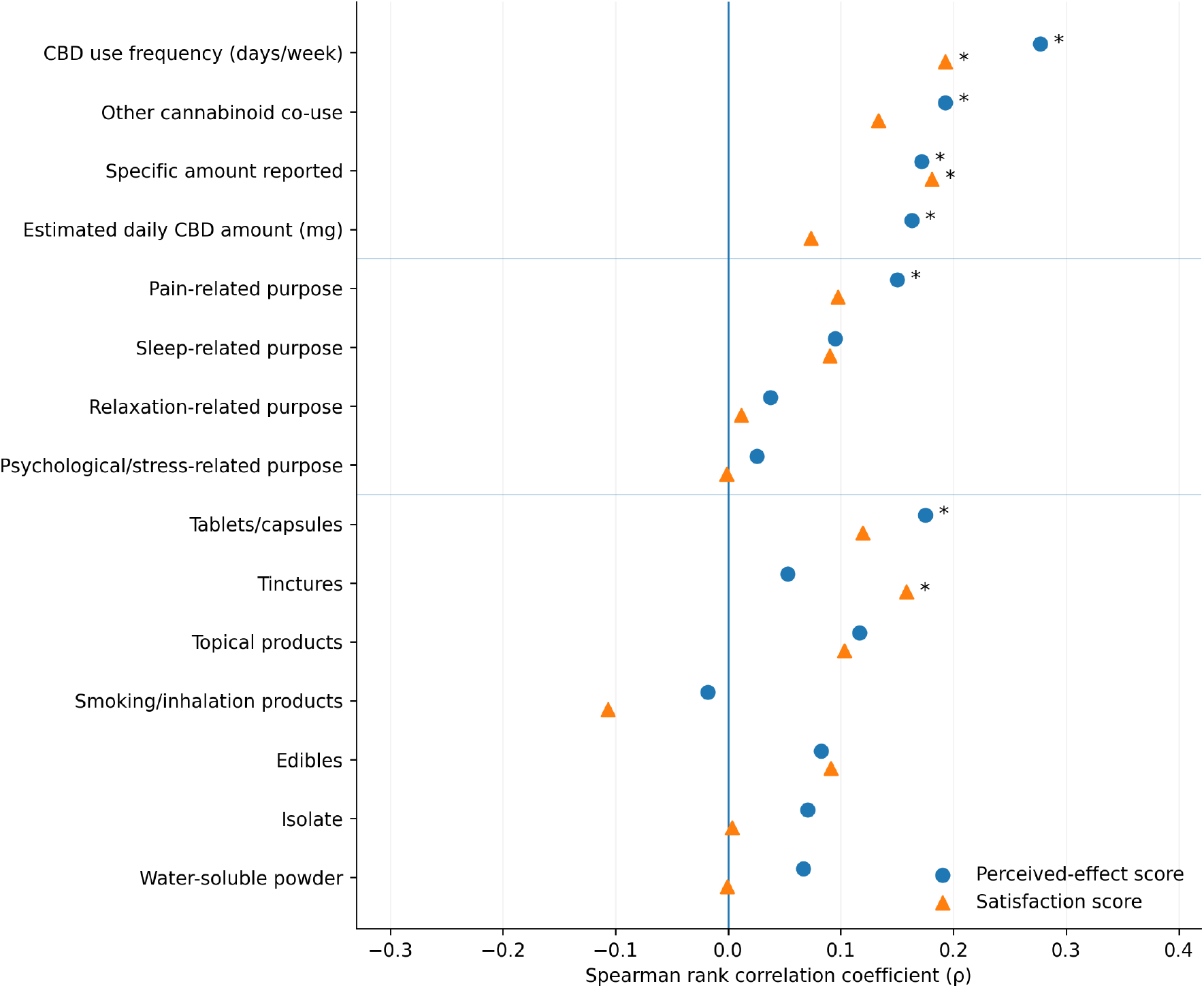
CBD use frequency and specific CBD amount-reporting status were the only characteristics correlated with both perceived effects and satisfaction. Spearman rank correlations of use characteristics with perceived-effect (circles) and satisfaction (triangles) scores. Variables are grouped by use status, purposes of use, and product form, and ordered within groups by the larger absolute correlation. *FDR-adjusted p < 0.05 (Benjamini–Hochberg). N = 373/377 (perceived effect/satisfaction), except estimated daily CBD amount (265/268) and co-use of other cannabinoids (301/304).

### Use patterns and sleep timing

Hierarchical clustering of 15 use characteristics yielded two clusters (Silhouette coefficient 0.219) separated mainly by use frequency (median 7.0 vs 1.5 days/week). In PCA, a component centered on amount-reporting status and co-use of other cannabinoids was preserved across two preprocessing approaches (r = 0.946 between corresponding component scores), whereas the frequency-dominated component was not (Supplementary Tables S8 and S9; Supplementary Figures S1 and S2).

Neither MSFsc nor SJL showed a clear adjusted association with perceived-effect score(MSFsc: B = −0.051, 95% CI −0.122 to 0.021; SJL: B = −0.053, 95% CI −0.186 to 0.079) or satisfaction score (MSFsc: B = −0.063, 95% CI −0.143 to 0.016; SJL: B = −0.060, 95% CI−0.209 to 0.089) after adjustment (Supplementary Table S11). No comparison of MSFsc by any of the 12 individual purposes of use, including the three sleep-related purposes, remained significant after FDR correction (Supplementary Table S10). These results suggest that use characteristics were structured mainly around use frequency and amount-reporting status, and that sleep timing was associated neither with subjective evaluation after adjustment nor with purposes of use.

## Discussion

In this study, use frequency was consistently associated with both perceived effects and satisfaction. Estimated daily amount was not clearly associated with either outcome after adjustment, whereas whether a specific milligram amount could be reported was associated with both, and amount-reporting status was in turn related to product form.

The association with use frequency persisted across the categorical sensitivity analysis and all supplementary models, although the models explained little of the between-person variation (adjusted R-squared 0.074 and 0.043). Regular use may allow users to recognize effects more readily; equally, satisfied users may continue or increase their use, so this association cannot be read as evidence that more frequent use improves perceived effects or satisfaction.

The more distinctive finding is the contrast between how much CBD participants reported using and whether they could report it at all. The weak unadjusted correlation between estimated daily amount and perceived effects did not persist after adjustment, and even at the upper confidence limit, a difference spanning the interquartile range of reported amount (5 to 38 mg) would correspond to about 0.25 points in perceived-effect score, less than half the difference associated with amount-reporting status (0.54 points). Reported amounts may also diverge from actual exposure, because labeled and measured CBD content can differ in commercial products (Bonn-Miller et al., 2017) and CBD absorption varies with route and food intake (Millar et al., 2018).

One possible explanation for this association is that respondents who can report a specific milligram amount may also track how they respond to it, which would allow them to reproduce a satisfactory amount or compare responses after changing it. Such tracking appears to be uncommon: in a previous Japanese survey, fewer than one-third of respondents with CBD use experience reported a daily CBD amount (Masataka et al., 2022).

Product form may help contextualize these findings. Some tincture products allow doses to be quantified using labeled concentrations and measuring devices, whereas quantifying amilligram amount from smoking or inhalation products may be more difficult. Consistent with this possibility, tincture use was more common and smoking or inhalation product use less common among respondents who reported specific milligram information. In PCA, amount-reporting status, other-cannabinoid co-use, and tincture use were positively correlated with a similar principal-component axis across the two preprocessing approaches, suggesting a possible broader pattern of product choice and use.

The two reporting groups did not differ significantly in age, sex, BMI, use frequency, or MSFsc. Reverse causation remains possible, in that users who perceive stronger effects may attend more closely to how much they take. Health literacy, which is associated with understanding of medication information and with medication self-management (Kim et al., 2022; Masumoto et al., 2023; Persell et al., 2020), and expectation, which contributes to subjective responses to cannabinoids (Gedin et al., 2022), are further candidates that future studies should measure directly.

Psychological or stress-related, relaxation-related, and sleep-related purposes were the most common, as in previous reports (Masataka et al., 2022; Moltke and Hindocha, 2021), yet none of the four purpose categories was associated with either outcome after adjustment. Unlike the motive-based profiles identified among people who co-used cannabis and CBD (Fortin et al., 2025), clustering in the present sample, which differed in population and in the characteristics analyzed, separated users mainly by use frequency, with modest separation (Silhouette coefficient 0.219).

Neither MSFsc nor SJL showed clear adjusted associations with either outcome, and none of the 12 comparisons of MSFsc by purpose of use, including sleep-related purposes, remained significant after FDR correction. These findings concern how CBD use is characterized rather than whether CBD improves sleep, on which evidence remains inconsistent: a recent meta-analysis reported improvements in subjective sleep outcomes for cannabinoids overall, whereas CBD-only interventions showed no clear benefit over placebo (da Silva et al., 2025).

Because the data were cross-sectional and self-reported, and both outcomes were single-item measures, the associations observed here cannot establish direction, and this study cannot identify an appropriate dose for commercially available CBD products. Participants were recruited through a CBD-related event, retail stores, and social media and may not represent all CBD users in Japan. The regulatory context has also shifted since the survey: CBN, named as an example in the survey item on other cannabinoids, was listed as a Designated Substance in June 2026 (Ministry of Health, Labour and Welfare, 2026). Nevertheless, the findings indicate that characterizing real-world CBD use may benefit from considering not only reported amount but also specific amount-reporting status and the associated product forms.

Prospective studies should record product concentration, amount per administration, dose adjustment, and subjective response over time.

## Conclusion

Subjective evaluations of CBD use in Japan were more consistently associated with use frequency and specific CBD amount-reporting status than with the reported daily amount. Amount-reporting status was also associated with product form, suggesting that whether respondents provided specific milligram information may be a useful dimension for characterizing real-world CBD use.

## Data Availability

All data produced in the present study are available upon reasonable request to the authors

## Author Contributions

AK: formal analysis, writing (original draft). CP: questionnaire design, data collection, formal analysis (early stage). LN: formal analysis (early-stage support). CN: conceptualization, methodology, supervision, project administration, formal analysis, writing (review and editing). All authors approved the final manuscript.

## Data Availability

The data supporting the findings of this study are available from the corresponding author upon reasonable request.

