## Supplementary materials for "CBD use characteristics associated with perceived effects and satisfaction among CBD product users in Japan: an exploratory analysis of a cross-sectional web-based survey"

*Correspondence: Chihiro Nozaki

#### Supplementary Methods

##### Study design and analytic sample

This was a cross-sectional analysis of an anonymous web-based survey of CBD product users in Japan conducted by our group between 24 November 2022 and 5 January 2023. We administered the survey using Google Forms as an open survey, and participation was voluntary and uncompensated. Each Google account could submit only one response; account information, including email addresses, was not collected, so responses remained anonymous to the researchers.

Eligible participants were respondents who were using CBD products at the time of the survey, and eligibility was assessed by reviewing each respondent's answers as a whole. Among the 382 respondents, four answered “never used,” “0,” “0 mg,” or “not using” to the item asking, “On average, how many mg of CBD do you take per day?” Three of these also indicated that they had not used CBD when asked about perceived effects, and the fourth entered “.” for perceived effects, which could not be interpreted; current CBD use therefore could not be confirmed for any of the four. One additional respondent stated in a free-text response that their answers should not be included in statistical analyses. These five respondents were excluded, leaving 377 participants in the final analytic sample. No a priori sample-size calculation was performed; all eligible respondents were included.

##### Measures

Survey items and response options are listed in Supplementary Table S1, and free-text responses were coded using decision rules finalized before analysis (Supplementary Table S2).

Original age bands were combined into four categories (<25, 25–34, 35–44, and ≥45 years), with ≥45 years as the reference category in regression analyses. Sex was coded male = 0 (reference) and female = 1; the one respondent who selected “neither” was retained in descriptive analyses and treated as missing in analyses requiring a binary covariate. BMI was calculated from self-reported height and weight.

Options of the multiple-response item “What effects do you expect when purchasing CBD?” were grouped into four binary purpose-of-use categories, each coded 1 if any of its options was selected: sleep-related (“help me fall asleep,” “deepen sleep,” “improve awakening”), psychological/stress-related (“reduce anxiety,” “reduce stress,” “calm me down”), pain-related (“reduce pain”), and relaxation-related (“relaxation effect”). Other options were not used. Seven binary product-form variables were created from the multiple-response item “CBD products you commonly purchase”: tinctures, tablets/capsules, smoking/inhalation products, edibles, topical products, isolate, and water-soluble powder. Free-text responses were assigned to a form only when a dosage form could be identified.

Use frequency was converted to 0.5, 1.5, 3.5, 5.5, and 7.0 days/week and analyzed as a continuous variable. Estimated daily CBD amount was converted to representative values of 5, 18, 38, 63, 88, 125, 175, and 225 mg, with the open-ended ≥201-mg category treated as 201–250 mg. Specific CBD amount-reporting status was coded 1 when a predefined category or a specific milligram amount was given, and 0 for “do not know,” blank responses, or responses without a determinable milligram amount (e.g., puff counts or product concentration only). Two respondents whose milligram information could not be converted to a daily average were coded 1, with estimated daily amount treated as missing.

Co-use of other cannabinoids was classified as no co-use, co-use, or unknown. Unknown status was treated as missing in regression and Spearman analyses and was represented in clustering by two binary variables (co-use and unknown).

Perceived effects were scored from “not at all” (0) to “very much” (5) and satisfaction from “very dissatisfied” (0) to “very satisfied” (6); free-text perceived-effect responses that could not be mapped to a response level were treated as missing.

The usual (e.g., weekday) schedule and the day-off schedule were used as workday and free-day inputs, respectively, to derive MSFsc and SJL; definitions, calculations, and the data-cleaning rule are given in Supplementary Table S3.

**Statistical analysis**

***Descriptive statistics***

Categorical variables were summarized as counts and percentages, and continuous variables as means, standard deviations, medians, and quartiles. Percentages for multiple-response items used the full analytic sample as the denominator, and estimated daily amount was summarized among the 268 participants with a quantifiable amount.

***Regression analyses***

Perceived-effect and satisfaction scores were analyzed separately by OLS regression, with the ordered scores treated as continuous. The main model included CBD use frequency, age, sex, and BMI; use frequency served as the primary explanatory variable because it was available for all participants, whereas estimated daily amount could not be quantified for 109. Supplementary models added estimated daily amount and co-use of other cannabinoids (model A), the four purpose-of-use categories (model B), or amount-reporting status (model C). All models used complete cases and HC3 robust standard errors, and results are reported as unstandardized coefficients (B) with 95% CIs, p values, $R^2$, and adjusted $R^2$.

Two sensitivity analyses were conducted. Use frequency was entered as its five original categories (reference, <1 day/week), with the overall association tested by a Wald F test based on the HC3 variance–covariance matrix, and the representative value for the ≥201-mg category in model A was varied from 225 mg to 201 mg and 250 mg.

For the main and supplementary models, residual-versus-fitted and Q–Q plots were inspected, and diagnostics were assessed using the Koenker–Breusch–Pagan test, variance inflation factors, Cook's distance, leverage, and externally studentized residuals. Heteroscedasticity was detected in all four satisfaction models (Koenker–Breusch–Pagan $p < 0.05$), supporting the use of HC3 robust standard errors; all variance inflation factors were below 2.0, and no observation had an absolute externally studentized residual greater than 3.

***Comparisons by amount-reporting status***

Participants who did and did not report a specific amount were compared on age and sex using Pearson's chi-square test with Cramér's V, and on BMI, use frequency, MSFsc, and both outcome scores using the Mann–Whitney U test with rank-biserial correlation. Each of the seven product forms was compared using the chi-square test, or Fisher's exact test for small expected counts, with odds ratios and 95% CIs; p values across the seven comparisons were adjusted using the Benjamini–Hochberg false discovery rate (FDR) procedure.

***Spearman correlations***

Spearman correlations were calculated pairwise among 17 variables: four use-status variables, four purpose-of-use categories, seven product forms, and both outcome scores. Of the 136 possible pairs, estimated daily amount × amount-reporting status was excluded because one is derived from the other, and p values for the remaining 135 pairs were FDR-adjusted.

***Hierarchical clustering***

Ward's hierarchical clustering with Euclidean distance was applied to 15 use characteristics: use frequency, amount-reporting status, co-use (two binary variables), four purpose-of-use categories, and seven product forms. Amount-reporting status was used in place of estimated daily amount to retain all 377 participants. Use frequency was standardized, whereas binary variables were kept as 0/1 so that rare characteristics would not be overweighted; outcome scores were not used. Solutions with 2–8 clusters were compared using the Silhouette coefficient, Calinski–Harabasz index, and Davies–Bouldin index, together with cluster size and interpretability. Cluster-forming variables were described without significance testing, and outcomes, age, sex, and BMI were compared between clusters using the tests described above.

##### *Principal component analysis*

PCA was applied to the same 15 variables with the same preprocessing to visualize the clustering feature space. Scores on the first two components were plotted by cluster, and their Pearson correlations with each original variable were calculated. As a sensitivity analysis, PCA was repeated after standardizing all 15 variables.

##### *Sleep-timing analyses*

MSFsc and SJL were each entered into OLS models for each outcome, adjusted for use frequency, amount-reporting status, age, sex, and BMI, with HC3 robust standard errors. MSFsc was also compared between participants who did and did not select each of the 12 individual purposes of use using the Mann–Whitney U test, with FDR adjustment across the 12 comparisons.

##### *Software and significance*

### Supplementary Figures

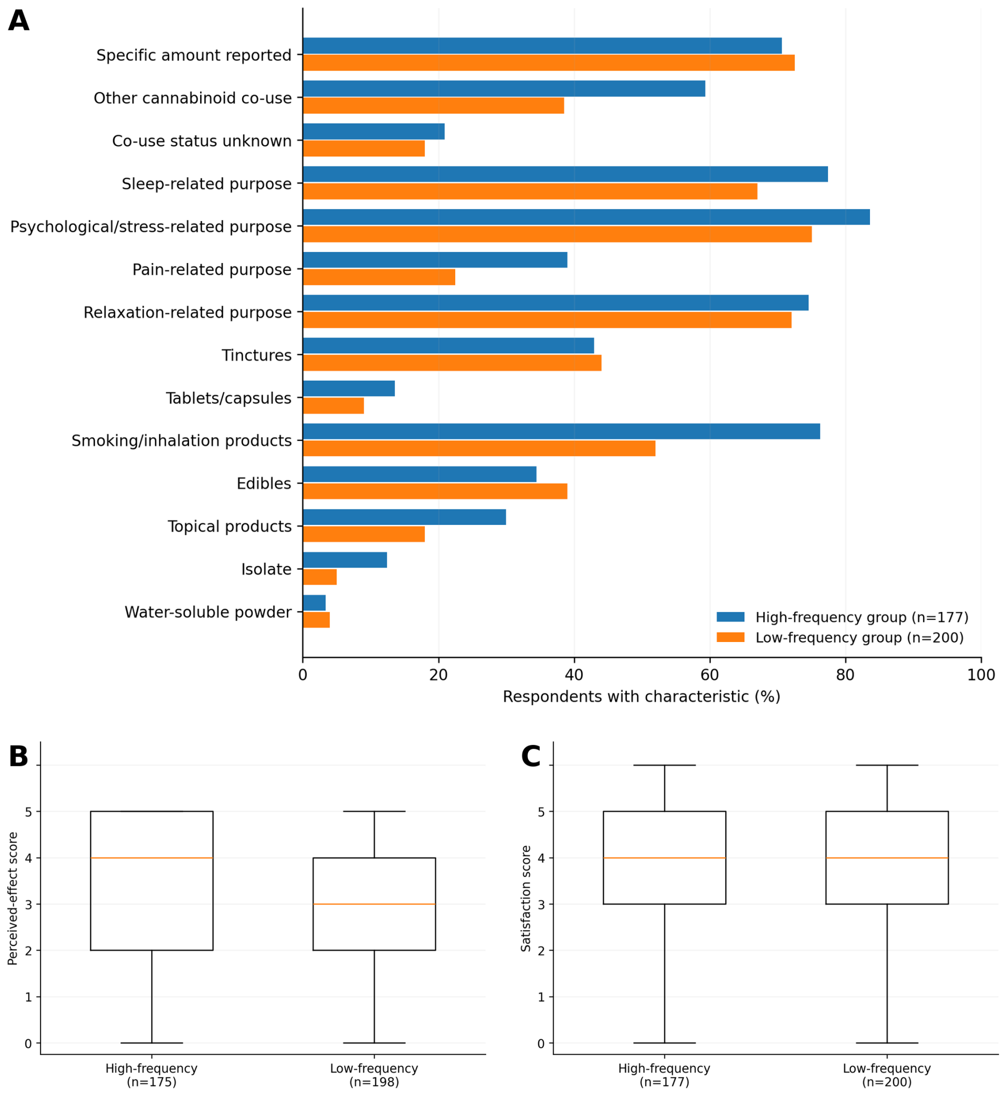


**Supplementary Figure S1.** The high-frequency-use group showed higher rank distributions of perceived effects and satisfaction than the low-frequency-use group.

A: Distribution of cluster-forming use characteristics. Binary variables are shown as the proportion of participants in each cluster with the characteristic; no significance tests were performed for these variables. B: Distribution of perceived-effect score. C: Distribution of satisfaction score. Boxes indicate the first quartile, median, and third quartile. Outcomes were compared using the Mann–Whitney U test.

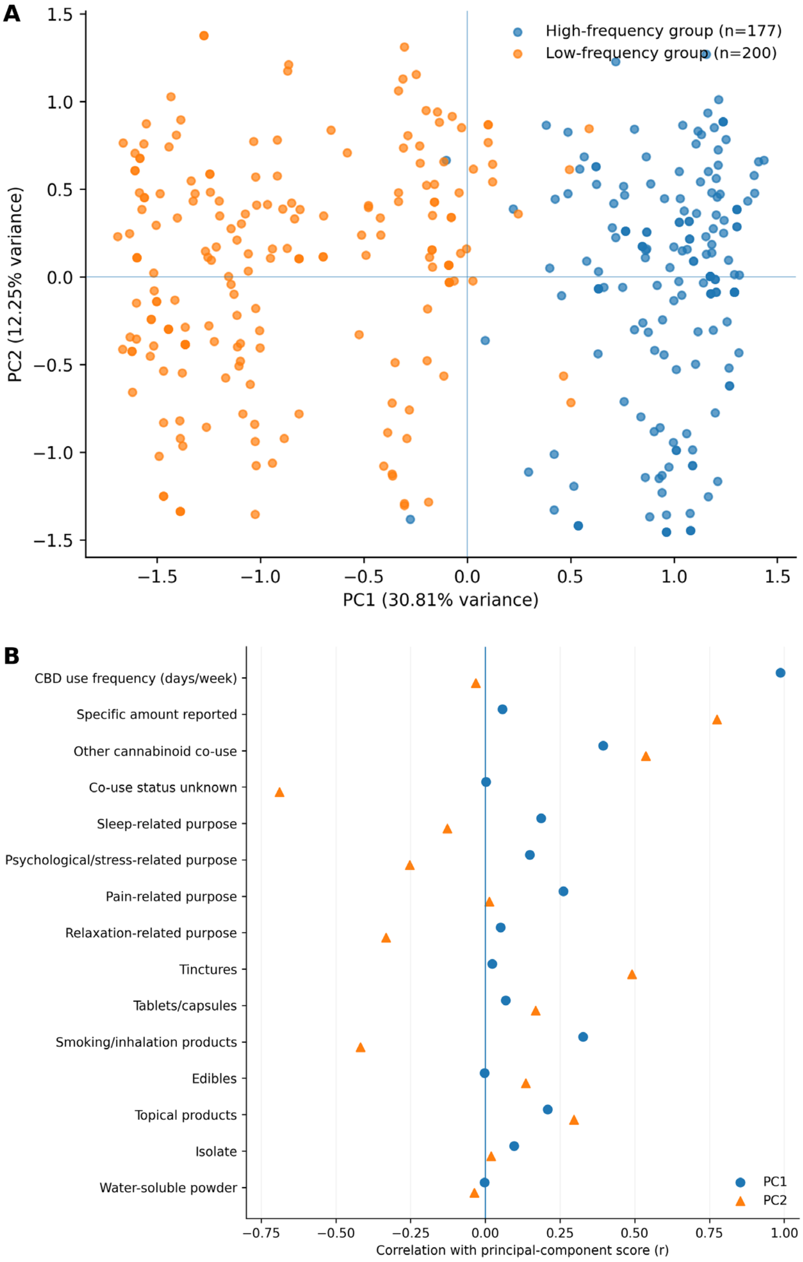


**Supplementary Figure S2.** The high- and low-frequency-use groups differed mainly along PC1, which primarily reflected CBD use frequency under the primary preprocessing, but the groups overlapped.

A: Score plot of participant positions on PC1 and PC2 for the 15 variables used in clustering, stratified by cluster. Each point represents one participant. B: Correlations between the original variables and PC1 and PC2 scores. PC1 was very strongly positively correlated with CBD use frequency (r = 0.988). PC1 and PC2 explained 30.81% and 12.25% of the variance, respectively (cumulative 43.06%).

#### Supplementary Tables

**Supplementary Table S1.** Survey questions and response options used in this study

| # | Measure | Question | Response options |
|---|---|---|---|
| 1 | Age | Age group | ≤19 years; 20–24; 25–29; 30–34; 35–39; 40–44; 45–49; 50–54; 55–59; 60–64; 65–69; ≥70 years |
| 2 | Sex | Biological sex | Male; female; neither |
| 3 | Purposes of use | What effects do you expect when purchasing CBD? (multiple responses allowed) | Help me fall asleep; deepen sleep; improve awakening; reduce anxiety; reduce stress; reduce fatigue; reduce pain; relaxation effect; clear my head; calm me down; reduce inflammation (e.g., skin irritation); allergy countermeasures; other (free text) |
| 4 | Product form | CBD products you commonly purchase (multiple responses allowed) | Tincture (oral oil); tablets/capsules (e.g., oil capsules); smoking/inhalation products (e.g., joints or vapes); edibles (e.g., gummies or cookies); topical products (e.g., balms or creams); isolate; water-soluble powder; other (free text) |
| 5 | Use frequency | How many days per week do you take CBD? | <1 day; 1–2 days; 3–4 days; 5–6 days; every day |
| 6 | Estimated daily CBD amount | On average, how many mg of CBD do you take per day? | 1–10 mg; 11–25 mg; 26–50 mg; 51–75 mg; 76–100 mg; 101–150 mg; 151–200 mg; ≥201 mg; do not know |
| 7 | Co-use of cannabinoids other than CBD | On average, how many mg per day in total do you take of cannabinoids other than CBD (e.g., CBN, CBG)? | I do not take any; 1–10 mg; 11–25 mg; 26–50 mg; 51–75 mg; 76–100 mg; 101–150 mg; 151–200 mg; ≥201 mg; do not know; other (free text) |
| 8 | Perceived effect | Do you feel the effects you expected? | Very much; moderately; slightly; sometimes yes and sometimes no; not much; not at all; other (free text) |
| 9 | Satisfaction | Are you satisfied with your current CBD intake and effects? | Very satisfied; satisfied; slightly satisfied; neither satisfied nor dissatisfied; slightly dissatisfied; dissatisfied; very dissatisfied |
| 10 | Usual-day wake time | At approximately what time do you usually wake up (e.g., on weekdays)? | 24-hour time (drop-down) |
| 11 | Usual-day bedtime | At approximately what time do you usually go to bed (e.g., on weekdays)? | 24-hour time (drop-down) |
| 12 | Day-off wake time | What time do you wake up on days off when you have nothing scheduled? | 24-hour time (drop-down) |
| 13 | Day-off bedtime | What time do you go to bed when the following day is a day off with | 24-hour time (drop-down) |

| | | | |
|---|---|---|---|
| | | nothing scheduled? | |
| 14 | Height | Height [cm] | Numeric (cm) |
| 15 | Weight | Weight [kg] | Numeric (kg) |

CBD, cannabidiol; CBN, cannabinol; CBG, cannabigerol.

**Supplementary Table S2.** Coding rules and analytic handling of free-text responses

| Variable | Free-text response | Analytic classification | Coded value | Missing-data handling | Reason |
|---|---|---|---|---|---|
| Purpose of use | I do not really understand CBD | Not in four categories | All four variables = 0 | Not missing | No corresponding purpose within the four categories could be identified, but a response was present |
| Purpose of use | Relief of hangover headache | Not in four categories | All four variables = 0 | Not missing | Free-text responses were not added to the four categories; categories were created from standard response options only |
| Product form | Cookie | Edible | Edible = 1 | Not missing | Product form could be clearly identified |
| Product form | Treatment oil | Topical product | Topical product = 1 | Not missing | Could be identified as a topical product |
| Product form | Full spectrum | Not in seven forms | All seven variables = 0 | Not missing | Describes composition rather than product form |
| Product form | Sample only | Not in seven forms | All seven variables = 0 | Not missing | Use was confirmed, but product form could not be identified |
| Estimated daily CBD amount | Less than 1 mg once per month | Specific amount reported | Amount reporting = 1; estimated daily amount = missing | Amount only missing | Milligram information was provided, but it could not be converted to an average daily amount |
| Estimated daily CBD amount | 50 to 70, sometimes 150 | Specific amount reported | Amount reporting = 1; estimated daily amount = missing | Amount only missing | Usual and maximum amounts were mixed, preventing determination of an average amount |
| Estimated daily CBD amount | About 5 puffs per day | No specific amount reported | Amount reporting = 0; estimated daily amount = missing | Amount missing | Milligrams per puff were unknown |

| | | | | | |
|---|---|---|---|---|---|
| Perceived effect | Only my second day | Cannot classify on scale | Perceived-effect score = missing | Missing | Degree of perceived effect could not be mapped to the 0–5 scale |

**Supplementary Table S3.** Definitions and calculation of sleep-timing-related indicators

| Indicator | Definition | Calculation | Role in this study |
|---|---|---|---|
| MSW | Mid-sleep on workdays | Workday bedtime + workday sleep duration / 2 | Workday sleep timing |
| MSF | Mid-sleep on free days | Free-day bedtime + free-day sleep duration / 2 | Free-day sleep timing |
| SDweek | Average weekly sleep duration | (Workday sleep duration × 5 + free-day sleep duration × 2) / 7 | Used to correct MSFsc |
| MSFsc | MSF corrected for oversleep on free days associated with sleep debt | If free-day sleep duration > SDweek: MSF − (free-day sleep duration − SDweek) / 2; otherwise MSF | Chronotype-related indicator following the MCTQ calculation approach |
| SJL | Difference between workday and free-day mid-sleep | Shortest absolute difference on a 24-hour cycle between MSF and MSW | Indicator related to mismatch between social time and sleep timing |

Sleep duration was calculated with wake times earlier than bedtime assigned to the following day. When the duration fell outside 3–14 hours, bedtime was shifted 12 hours later to correct a possible 12-hour-format entry error (23 usual-day and 29 day-off bedtimes corrected); responses still outside this range, or with identical bedtime and wake time, were excluded from sleep-timing analyses. Daytime sleep schedules yielding 3–14 hours were retained. MSW, mid-sleep on workdays; MSF, mid-sleep on free days; SDweek, average weekly sleep duration; MSFsc, sleep-corrected MSF; SJL, social jetlag; MCTQ, Munich ChronoType Questionnaire.

**Supplementary Table S4.** Regression coefficients for the main and supplementary models

| Predictor | Main model | Model A | Model B | Model C |
|---|---|---|---|---|
| *A. Perceived-effect score* | | | | |
| CBD use frequency (days/week) | 0.148*** (0.091 to 0.205) | 0.126*** (0.053 to 0.200) | 0.137*** (0.079 to 0.195) | 0.145*** (0.089 to 0.201) |
| Estimated daily CBD amount (mg) | – | 0.0035 (−0.0004 to 0.0075) | – | – |
| Co-use of other cannabinoids | – | 0.093 (−0.258 to 0.444) | – | – |
| Sleep-related purpose | – | – | 0.169 (−0.172 to 0.511) | – |
| Psychological/stress-related purpose | – | – | −0.079 (−0.463 to 0.306) | – |
| Pain-related purpose | – | – | 0.257 (−0.053 to 0.567) | – |

| | | | | |
|---|---|---|---|---|
| Relaxation-related purpose | – | – | 0.087 (−0.259 to 0.433) | – |
| Specific amount reported | – | – | – | 0.543*** (0.223 to 0.864) |
| Age <25 years | 0.244 (−0.278 to 0.766) | 0.102 (−0.512 to 0.716) | 0.263 (−0.268 to 0.793) | 0.232 (−0.284 to 0.748) |
| Age 25–34 years | 0.048 (−0.353 to 0.448) | −0.023 (−0.462 to 0.415) | 0.053 (−0.351 to 0.456) | 0.056 (−0.336 to 0.447) |
| Age 35–44 years | 0.290 (−0.098 to 0.677) | 0.263 (−0.180 to 0.707) | 0.262 (−0.130 to 0.654) | 0.295 (−0.086 to 0.675) |
| Female | 0.261 (−0.027 to 0.550) | 0.394* (0.056 to 0.732) | 0.232 (−0.072 to 0.536) | 0.273 (−0.011 to 0.558) |
| BMI (kg/m$^2$) | −0.007 (−0.039 to 0.025) | −0.021 (−0.055 to 0.013) | −0.008 (−0.041 to 0.025) | −0.009 (−0.041 to 0.023) |
| n | 372 | 253 | 372 | 372 |
| $R^2$ | 0.089 | 0.111 | 0.101 | 0.120 |
| Adjusted $R^2$ | 0.074 | 0.082 | 0.076 | 0.103 |
| *B. Satisfaction score* | | | | |
| CBD use frequency (days/week) | 0.113*** (0.049 to 0.177) | 0.111* (0.024 to 0.197) | 0.107** (0.040 to 0.174) | 0.111*** (0.047 to 0.174) |
| Estimated daily CBD amount (mg) | – | −0.0013 (−0.0070 to 0.0045) | – | – |
| Co-use of other cannabinoids | – | 0.053 (−0.365 to 0.470) | – | – |
| Sleep-related purpose | – | – | 0.231 (−0.136 to 0.598) | – |
| Psychological/stress-related purpose | – | – | −0.124 (−0.533 to 0.284) | – |
| Pain-related purpose | – | – | 0.137 (−0.213 to 0.487) | – |
| Relaxation-related purpose | – | – | −0.076 (−0.436 to 0.285) | – |
| Specific amount reported | – | – | – | 0.559** (0.214 to 0.903) |
| Age <25 years | −0.049 (−0.658 to 0.560) | −0.343 (−1.047 to 0.361) | −0.044 (−0.655 to 0.567) | −0.062 (−0.661 to 0.537) |
| Age 25–34 years | −0.010 (−0.482 to 0.462) | −0.245 (−0.793 to 0.303) | −0.016 (−0.490 to 0.458) | −0.005 (−0.461 to 0.452) |
| Age 35–44 years | 0.338 (−0.100 to 0.777) | 0.296 (−0.206 to 0.798) | 0.304 (−0.137 to 0.746) | 0.347 (−0.076 to 0.771) |
| Female | 0.321* (0.001 to 0.640) | 0.424* (0.037 to 0.811) | 0.312 (−0.017 to 0.640) | 0.329* (0.014 to 0.645) |
| BMI (kg/m$^2$) | −0.013 (−0.051 to 0.025) | −0.034 (−0.076 to 0.009) | −0.015 (−0.054 to 0.024) | −0.015 (−0.053 to 0.023) |
| n | 376 | 256 | 376 | 376 |
| $R^2$ | 0.059 | 0.089 | 0.066 | 0.085 |
| Adjusted $R^2$ | 0.043 | 0.059 | 0.041 | 0.067 |

Values are unstandardized regression coefficients (B) with 95% confidence intervals based on HC3 robust standard errors. Model A added estimated daily CBD amount and co-use of other cannabinoids, model B the four purpose-of-use categories, and model C specific CBD amount-reporting status to the main model. Reference categories: age ≥45 years; male; no co-use; no specific amount reported. Assigning 201 or 250 mg instead of 225 mg to the ≥201-mg category changed the coefficient for estimated daily amount by no more than 0.0001 for either outcome (all $p > 0.05$). *$p < 0.05$; **$p < 0.01$; ***$p < 0.001$.

**Supplementary Table S5.** Sensitivity analysis treating CBD use frequency as five categories

| Predictor | Perceived-effect score | | | Satisfaction score | | |
|---|---|---|---|---|---|---|
| | **B** | **95% CI** | **p value** | **B** | **95% CI** | **p value** |
| Use frequency 1–2 days/week | 0.474 | −0.028 to 0.976 | 0.064 | 0.643 | 0.061 to 1.224 | 0.030 |
| Use frequency 3–4 days/week | 0.760 | 0.270 to 1.250 | 0.002 | 0.848 | 0.338 to 1.358 | 0.001 |
| Use frequency 5–6 days/week | 0.846 | 0.301 to 1.391 | 0.002 | 0.897 | 0.316 to 1.477 | 0.003 |
| Use frequency every day | 1.102 | 0.666 to 1.539 | <0.001 | 0.937 | 0.461 to 1.414 | <0.001 |
| Age: <25 years | 0.252 | −0.271 to 0.774 | 0.344 | −0.039 | −0.652 to 0.574 | 0.901 |
| Age: 25–34 years | 0.048 | −0.350 to 0.446 | 0.811 | −0.008 | −0.473 to 0.457 | 0.974 |
| Age: 35–44 years | 0.269 | −0.121 to 0.659 | 0.176 | 0.311 | −0.130 to 0.752 | 0.166 |
| Female | 0.273 | −0.018 to 0.565 | 0.066 | 0.346 | 0.026 to 0.667 | 0.034 |
| BMI (kg/m²) | −0.006 | −0.038 to 0.026 | 0.702 | −0.011 | −0.048 to 0.027 | 0.578 |

Values are unstandardized regression coefficients (B), 95% confidence intervals (CI), and p values based on HC3 robust standard errors. Reference categories: <1 day/week; age ≥45 years; male. Perceived-effect model, n = 372, $R^2 = 0.096$, adjusted $R^2 = 0.074$; satisfaction model, n = 376, $R^2 = 0.074$, adjusted $R^2 = 0.051$. Overall Wald F test for frequency category: $F(4, 362) = 6.724$, $p < 0.001$ (perceived effects); $F(4, 366) = 4.238$, $p = 0.002$ (satisfaction).

**Supplementary Table S6.** Participant characteristics and outcomes by specific CBD amount-reporting status

| Variable | Category/statistic | Specific amount reported (n = 270) | No specific amount reported (n = 107) | Test statistic | p value | Effect size |
|---|---|---|---|---|---|---|
| Age | <25 years | 36 | 13 | $\chi^2 = 0.178$ | 0.981 | Cramér's V = 0.022 |
| | 25–34 years | 104 | 43 | | | |
| | 35–44 years | 77 | 31 | | | |
| | ≥45 years | 53 | 20 | | | |
| Sex | Male | 161 | 61 | $\chi^2 = 0.256$ | 0.613 | Cramér's V = 0.026 |

| | | | | | | |
|---|---|---|---|---|---|---|
| | Female | 108 | 46 | | | |
| BMI (kg/m²) | Median [Q1–Q3] | 21.58 [19.61–24.88] (n = 270) | 21.51 [19.62–24.64] (n = 107) | U = 14712.0 | 0.780 | $r_{rb}$ = 0.018 |
| CBD use frequency (days/week) | Median [Q1–Q3] | 3.5 [1.5–7.0] (n = 270) | 3.5 [1.0–7.0] (n = 107) | U = 14982.0 | 0.560 | $r_{rb}$ = 0.037 |
| Perceived-effect score | Median [Q1–Q3] | 4.0 [2.0–4.5] (n = 267) | 3.0 [2.0–4.0] (n = 106) | U = 17168.5 | <0.001 | $r_{rb}$ = 0.213 |
| Satisfaction score | Median [Q1–Q3] | 4.0 [3.0–5.0] (n = 270) | 3.0 [3.0–5.0] (n = 107) | U = 17717.5 | <0.001 | $r_{rb}$ = 0.227 |
| MSFsc (h) | Median [Q1–Q3] | 4.79 [3.64–6.14] (n = 269) | 5.08 [3.90–6.25] (n = 104) | U = 13095.5 | 0.339 | $r_{rb}$ = −0.064 |

Values for age and sex are n. Age and sex were compared using Pearson's chi-square test with Cramér's V, excluding the one respondent who selected “neither” from the sex comparison; other variables were compared using the Mann–Whitney U test with rank-biserial correlation ($r_{rb}$). Q1, first quartile; Q3, third quartile; MSFsc, sleep-corrected mid-sleep on free days.

**Supplementary Table S7.** Spearman rank correlations among use characteristics and outcomes

| Variable | 1 | 2 | 3 | 4 | 5 | 6 | 7 | 8 | 9 | 10 | 11 | 12 | 13 | 14 | 15 | 16 |
|---|---|---|---|---|---|---|---|---|---|---|---|---|---|---|---|---|
| 1. CBD use frequency | | | | | | | | | | | | | | | | |
| 2. Estimated daily CBD amount | .197* | | | | | | | | | | | | | | | |
| 3. Specific amount reported | .030 | NA | | | | | | | | | | | | | | |
| 4. Co-use of other cannabinoids | .390* | .353* | .207* | | | | | | | | | | | | | |
| 5. Sleep-related purpose | .121 | −.007 | −.053 | .156* | | | | | | | | | | | | |
| 6. Psychological/stress-related purpose | .109 | .019 | −.078 | .124 | .098 | | | | | | | | | | | |
| 7. Pain-related purpose | .191* | .070 | −.072 | .186* | .193* | .126* | | | | | | | | | | |
| 8. Relaxation-related purpose | .038 | −.232* | −.102 | −.035 | .101 | .262* | .046 | | | | | | | | | |
| 9. Tinctures | .030 | .074 | .161* | −.062 | −.094 | −.074 | .110 | −.134* | | | | | | | | |
| 10. Tablets/capsules | .060 | .100 | .073 | .001 | −.022 | −.046 | .042 | −.071 | .131* | | | | | | | |
| 11. Smoking/inhalation products | .247* | .097 | −.161* | .347* | .149* | .231* | .021 | .162* | −.311* | −.081 | | | | | | |
| 12. Edibles | −.029 | .166* | .066 | .204* | .087 | .123 | .059 | .040 | .006 | .061 | .067 | | | | | |
| 13. Topical products | .171* | .061 | .045 | .198* | .042 | −.082 | .287* | −.129* | .205* | .161* | −.122 | .054 | | | | |
| 14. Isolate | .088 | .119 | .002 | .043 | .063 | .016 | .090 | −.031 | −.056 | .074 | −.045 | −.016 | .032 | | | |
| 15. Water-soluble powder | .018 | −.019 | −.094 | −.033 | .060 | −.037 | .054 | −.071 | .026 | .020 | −.113 | .024 | .056 | .242* | | |
| 16. Perceived-effect score | .277* | .163* | .172* | .193* | .095 | .025 | .150* | .038 | .053 | .175* | −.018 | .083 | .117 | .071 | .067 | |
| 17. Satisfaction score | .193* | .074 | .181* | .134 | .090 | −.001 | .097 | .012 | .158* | .119 | −.106 | .091 | .103 | .003 | −.001 | .681* |

Values are Spearman rank correlation coefficients calculated with pairwise deletion (N = 257 to 377). *FDR-adjusted $p < 0.05$ (Benjamini–Hochberg procedure across 135 pairs). NA, not estimated because specific amount reporting was 1 for all participants with a quantified daily amount.

**Supplementary Table S8.** Characteristics of the two clusters identified by hierarchical clustering

| **Variable** | **Category/statistic** | **Cluster 1: high frequency (n = 177)** | **Cluster 2: low frequency (n = 200)** | **Test statistic** | **p value** | **Effect size** |
|---|---|---|---|---|---|---|
| *Cluster-forming characteristics* | | | | | | |
| CBD use frequency (days/week) | | 7.0 [7.0–7.0] | 1.5 [0.5–3.5] | | | |
| Specific amount reported | | 125 (70.6%) | 145 (72.5%) | | | |
| Other cannabinoid co-use | | 105 (59.3%) | 77 (38.5%) | | | |
| Other cannabinoid co-use unknown | | 37 (20.9%) | 36 (18.0%) | | | |
| Sleep-related purpose | | 137 (77.4%) | 134 (67.0%) | | | |
| Psychological/stress-related purpose | | 148 (83.6%) | 150 (75.0%) | | | |
| Pain-related purpose | | 69 (39.0%) | 45 (22.5%) | | | |
| Relaxation-related purpose | | 132 (74.6%) | 144 (72.0%) | | | |
| Tinctures | | 76 (42.9%) | 88 (44.0%) | | | |
| Tablets/capsules | | 24 (13.6%) | 18 (9.0%) | | | |
| Smoking/inhalation products | | 135 (76.3%) | 104 (52.0%) | | | |
| Edibles | | 61 (34.5%) | 78 (39.0%) | | | |
| Topical products | | 53 (29.9%) | 36 (18.0%) | | | |
| Isolate | | 22 (12.4%) | 10 (5.0%) | | | |
| Water-soluble powder | | 6 (3.4%) | 8 (4.0%) | | | |
| *Background characteristics and outcomes* | | | | | | |
| Age | <25 years | 19 (10.7%) | 30 (15.0%) | $\chi^2$ = 12.437 | 0.006 | Cramér's V = 0.182 |
| | 25–34 years | 56 (31.6%) | 91 (45.5%) | | | |
| | 35–44 years | 60 (33.9%) | 48 (24.0%) | | | |
| | ≥45 years | 42 (23.7%) | 31 (15.5%) | | | |
| Sex | Male | 115 (65.0%) | 107 (53.5%) | $\chi^2$ = 4.862 | 0.027 | Cramér's V = 0.114 |
| | Female | 62 (35.0%) | 92 (46.0%) | | | |
| | Neither | 0 (0.0%) | 1 (0.5%) | | | |
| BMI (kg/m²) | Median [Q1–Q3] | 21.83 [19.66–25.39] (n = 177) | 21.26 [19.55–24.34] (n = 200) | U = 19073.0 | 0.194 | $r_{rb}$ = 0.078 |

| | | | | | | |
|---|---|---|---|---|---|---|
| Perceived-effect score | Median [Q1–Q3] | 4.0 [2.0–5.0] (n = 175) | 3.0 [2.0–4.0] (n = 198) | U = 21387.0 | <0.001 | $r_{rb}$ = 0.234 |
| Satisfaction score | Median [Q1–Q3] | 4.0 [3.0–5.0] (n = 177) | 4.0 [3.0–5.0] (n = 200) | U = 20589.0 | 0.005 | $r_{rb}$ = 0.163 |

Values are n (%) or median [Q1–Q3]. Cluster-forming characteristics were not tested. Age and sex were compared using Pearson's chi-square test with Cramér's V, excluding the one respondent who selected "neither" from the sex comparison; BMI and outcomes were compared using the Mann–Whitney U test with rank-biserial correlation ($r_{rb}$). Among solutions with 2–8 clusters, the two-cluster solution had the highest Silhouette coefficient (0.219) and Calinski–Harabasz index (122.3) and the lowest Davies–Bouldin index (1.72). Q1, first quartile; Q3, third quartile.

**Supplementary Table S9.** Correlations of the 15 clustering variables with the first two principal components under the primary and all-variable-standardized preprocessing

| Variable | Primary PC1 r | Primary PC2 r | All-z PC1 r | All-z PC2 r |
|---|---|---|---|---|
| CBD use frequency | 0.988 | −0.032 | 0.172 | 0.511 |
| Specific amount reported | 0.057 | 0.775 | 0.805 | −0.057 |
| Other cannabinoid co-use | 0.394 | 0.536 | 0.684 | 0.483 |
| Other cannabinoid co-use unknown | 0.002 | −0.689 | −0.768 | −0.011 |
| Sleep-related purpose | 0.186 | −0.127 | −0.013 | 0.467 |
| Psychological/stress-related purpose | 0.149 | −0.253 | −0.167 | 0.533 |
| Pain-related purpose | 0.261 | 0.014 | 0.072 | 0.342 |
| Relaxation-related purpose | 0.051 | −0.333 | −0.245 | 0.401 |
| Tinctures | 0.023 | 0.490 | 0.336 | −0.340 |
| Tablets/capsules | 0.068 | 0.168 | 0.213 | −0.096 |
| Smoking/inhalation products | 0.326 | −0.418 | −0.201 | 0.682 |
| Edibles | −0.003 | 0.135 | 0.151 | 0.253 |
| Topical products | 0.208 | 0.297 | 0.307 | 0.020 |
| Isolate | 0.096 | 0.020 | 0.061 | 0.084 |
| Water-soluble powder | −0.003 | −0.037 | −0.103 | −0.097 |

Values are Pearson correlation coefficients between each original variable and the component scores. Primary, primary preprocessing; All-z, all 15 variables standardized to mean 0 and standard deviation 1. Primary PC2 and all-z PC1 scores were very strongly correlated (r = 0.946). PC1 and PC2 explained 30.81% and 12.25% of the variance under the primary preprocessing (cumulative 43.06%) and 14.39% and 12.98% under all-variable standardization (cumulative 27.37%).

**Supplementary Table S10.** Comparison of MSFsc between participants who did and did not select each of 12 individual purposes of CBD use

| Purpose of use | Unadjusted p | FDR-adjusted p |
|---|---|---|
| Help me fall asleep | 0.333 | 0.732 |
| Deepen sleep | 0.989 | 0.989 |
| Improve awakening | 0.334 | 0.732 |
| Reduce anxiety | 0.741 | 0.889 |
| Reduce stress | 0.620 | 0.889 |
| Reduce fatigue | 0.342 | 0.732 |
| Reduce pain | 0.015 | 0.185 |
| Relaxation effect | 0.914 | 0.989 |
| Clear my head | 0.101 | 0.606 |
| Calm me down | 0.566 | 0.889 |
| Reduce inflammation | 0.720 | 0.889 |
| Allergy countermeasures | 0.366 | 0.732 |

Each comparison used the Mann–Whitney U test, with p values adjusted across the 12 tests using the Benjamini–Hochberg false discovery rate (FDR) procedure. MSFsc, sleep-corrected mid-sleep on free days.

**Supplementary Table S11.** Adjusted associations of sleep-timing indicators with perceived effects and satisfaction

| Indicator | Outcome | N | B | 95% CI | p value | $R^2$ |
|---|---|---|---|---|---|---|
| MSFsc | Perceived-effect score | 368 | −0.051 | −0.122 to 0.021 | 0.163 | 0.136 |
| MSFsc | Satisfaction score | 372 | −0.063 | −0.143 to 0.016 | 0.118 | 0.102 |
| SJL | Perceived-effect score | 368 | −0.053 | −0.186 to 0.079 | 0.429 | 0.129 |
| SJL | Satisfaction score | 372 | −0.060 | −0.209 to 0.089 | 0.428 | 0.093 |

Values are unstandardized regression coefficients (B), 95% confidence intervals (CI), and p values based on HC3 robust standard errors. Each model adjusted for CBD use frequency, specific CBD amount-reporting status, age, sex, and BMI. MSFsc, sleep-corrected mid-sleep on free days; SJL, social jetlag.
